# A shared genetic signature for common chronic pain conditions and its impact on biopsychosocial traits

**DOI:** 10.1101/2022.03.13.22272317

**Authors:** Scott F. Farrell, Pik-Fang Kho, Mischa Lundberg, Adrián I. Campos, Miguel E. Rentería, Rutger M. J. de Zoete, Michele Sterling, Trung Thanh Ngo, Gabriel Cuéllar-Partida

**Affiliations:** RECOVER Injury Research Centre, The University of Queensland, Herston QLD, Australia; NHMRC Centre of Research Excellence: Better Health Outcomes for Compensable Injury, The University of Queensland, Herston QLD, Australia; Tess Cramond Pain & Research Centre, Royal Brisbane & Women’s Hospital; Surgical, Treatment & Rehabilitation Service (STARS), Herston QLD, Australia; Division of Cardiovascular Medicine, Department of Medicine, Stanford University School of Medicine, Stanford CA, USA; Molecular Cancer Epidemiology Laboratory, Department of Genetics & Computational Biology, QIMR Berghofer Medical Research Institute, Herston QLD, Australia; School of Biomedical Sciences, Queensland University of Technology, Brisbane QLD, Australia; Diamantina Institute, The University of Queensland & Translational Research Institute, Woolloongabba QLD, Australia; Transformational Bioinformatics, CSIRO Health & Biosecurity, North Ryde NSW, Australia; Institute for Molecular Bioscience, The University of Queensland, St Lucia QLD, Australia; Genetic Epidemiology Laboratory, Department of Genetics & Computational Biology, QIMR Berghofer Medical Research Institute, Herston QLD, Australia; School of Allied Health Science & Practice, The University of Adelaide, Adelaide SA, Australia

**Keywords:** Chronic Pain, Musculoskeletal Pain, Genome-Wide Association Studies, Genetic Predisposition to Disease, Precision Medicine

## Abstract

The multifactorial nature of chronic pain with its numerous comorbidities presents a formidable challenge in disentangling their aetiology. Here, we performed genome-wide association studies of eight regional chronic pain types using UK Biobank data (N=4,037–79,089 cases; N=239,125 controls), followed by bivariate linkage disequilibrium-score regression and latent causal variable analyses to determine (respectively) their genetic correlations and genetic causal proportion (GCP) parameters with 1,492 other complex traits. We report evidence of a shared genetic signature across common chronic pain types as their genetic correlations and GCP parameter directions were broadly consistent across a wide array of biopsychosocial traits. Across 5,942 significant genetic correlations, 570 trait pairs could be explained by a causal association (|GCP| > 0.6; 5% false discovery rate), including 82 traits affected by pain while 488 contributed to an increased risk of chronic pain such as certain somatic pathologies (e.g., musculoskeletal), psychiatric traits (e.g., depression), socioeconomic factors (e.g., occupation) and medical comorbidities (e.g., cardiovascular disease). This data-driven study has demonstrated a novel & efficient strategy for identifying genetically supported risk & protective traits to enhance the design of interventional trials targeting underlying causal factors and help accelerate the development of more effective treatments with broader clinical utility.

## Introduction

Despite enormous research efforts into determining the aetiology of chronic pain, common musculoskeletal conditions such as back pain and neck pain remain a major clinical and scientific challenge.^1^ Over the past decade however, large-scale genome-wide association studies (GWAS) have made inroads towards elucidating the biological pathways and complex traits contributing to a wide range of conditions such as neurological, psychiatric & cardiometabolic disorders.^2^ While such studies have provided evidence for genetically supported therapeutic targets & mechanisms,^3,4^ the application of statistical genetics methods to musculoskeletal^5–17^ and chronic pain conditions^18–25^ as a strategy towards disentangling their causal mechanisms requires further comprehensive & concerted effort.

Previous clinical & epidemiological studies have found a range of psychiatric (e.g., stress, depression, trauma, anxiety^26,27^), lifestyle (e.g., smoking,^28,29^ sedentary behaviour^30,31^) and socioeconomic factors (e.g., income & education levels^32,33^) associated with an increased risk of chronic pain, while medical conditions such as obesity^34^ & cardiovascular diseases^35,36^ are also common comorbidities.^37^ Large-scale GWAS have identified specific genetic loci in chronic pain conditions (e.g., back,^12,19^ neck/shoulder,^15^ knee pain^9^ & widespread pain^14,20^) and demonstrated shared genetic bases with biopsychosocial traits (e.g., depression,^19,20,27^ post-traumatic stress disorder,^20^ anorexia nervosa,^38^ asthma,^20^ brain morphology,^18^ poor sleep,^8,19,39^ obesity,^5^ low educational attainment^19,40^ & smoking^19,40^). However, the nature of causal relationships between common chronic pain conditions and such multidimensional factors remains to be comprehensively examined.

Here we investigated the genetic correlations and causal relationships between eight regional chronic pain types and a comprehensive range of biological, psychological/psychiatric & social traits. In the largest phenome-wide study of chronic pain to date, we leveraged GWAS summary statistics in the Complex Traits Genetics Virtual Lab (CTG-VL — http://genoma.io)^41^ to delineate the putative genetic causal directions across common chronic pain conditions and >1,400 other traits/disorders.

## Materials & Methods

### Discovery GWAS Datasets for Chronic Pain

An overview of the analysis pipeline employed in the present study is shown in Figure 1. We performed GWAS of regional chronic pain types using the UK Biobank resource (application number 25331). The UK Biobank holds research ethics approval from the North West Multi-centre Research Ethics Committee (Manchester, United Kingdom). All participants provided written informed consent.

**Figure 1:**
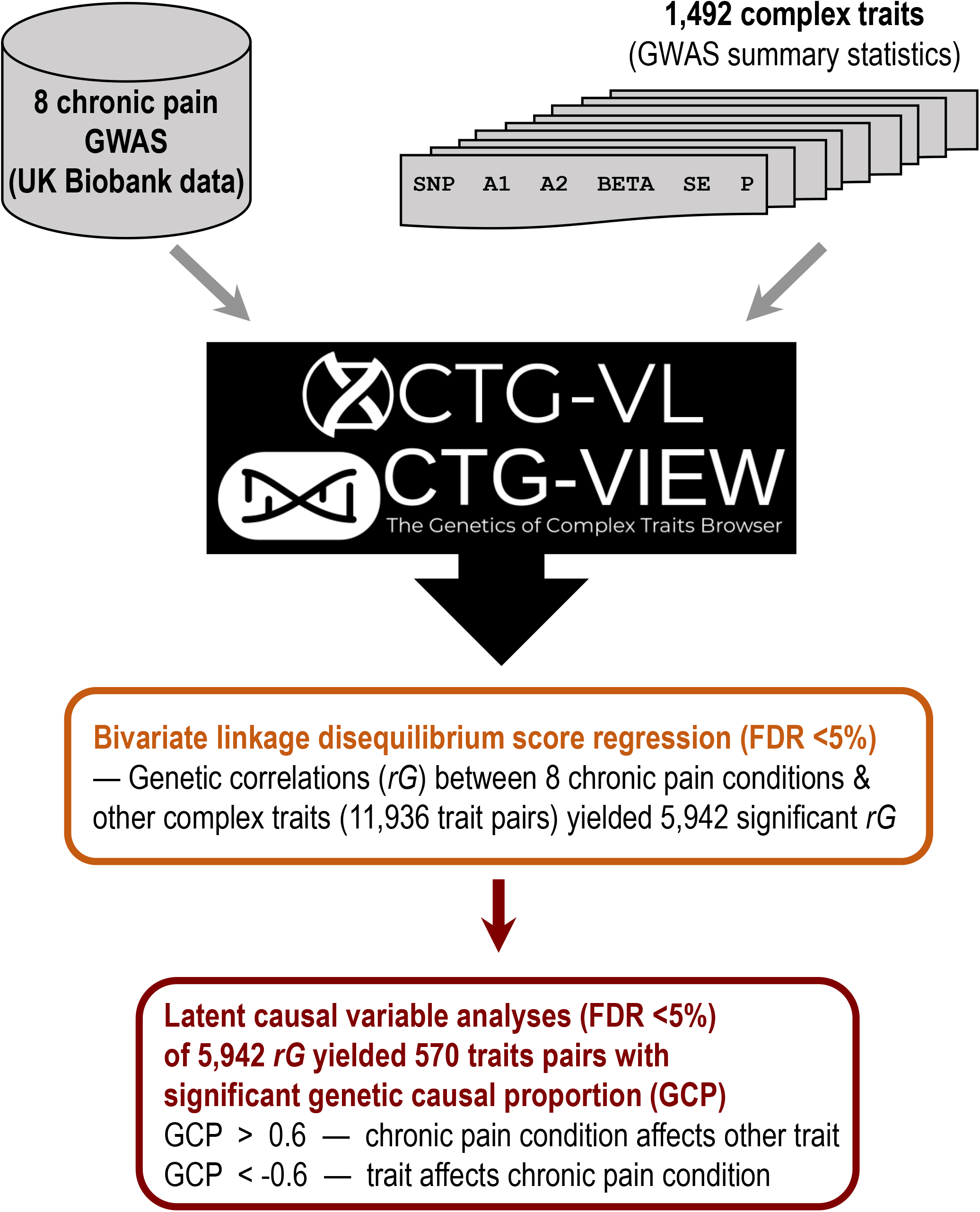
Phenome-wide analysis pipeline for eight types of regional chronic pain with 1,492 other complex traits. Genome-wide association studies (GWAS) were performed for chronic back, neck/shoulder, hip, knee, abdominal, facial, headache and widespread pain. These results and publicly available GWAS summary statistics for 1,492 complex traits were uploaded into the Complex Traits Genetics Virtual Lab (CTG-VL), which along with CTG-VIEW were used to perform cross-trait linkage disequilibrium-score (LD-score) regression and latent causal variable analyses to respectively calculate genetic correlations (*rG*) and genetic causal proportions (GCP). The 570 traits with significant GCPs were categorised as per Figures 4–7 (x-axis) and Supplementary Tables S2–S9. This strategy enables the identification & targeting of genetically supported risk & protective factors for chronic pain types in further translationally focused studies (e.g., interventional trials).

Chronic pain was defined using self-report answers to the following questions: *Have you had [back pains* / *neck or shoulder pains* / *hip pains* / *knee pains* / *abdominal pains* / *facial pains* / *headaches* / *pain all over the body] for more than 3 months?* (Questionnaire field ID: 6159). These questions could be answered with *Yes*, *No*, *Don’t know*, or *Prefer not to answer*. Participants who responded *Yes* to a question were defined as cases for that specific chronic pain type (i.e., chronic back pain, chronic widespread pain [pain all over the body] etc.). We defined controls as participants who denied experiencing any pain for more than three months (N=239,125). Participants who preferred not to answer were excluded from the study. The sample sizes of each GWAS were as follows: back pain (N=79,089); neck/shoulder pain (N=72,216); hip pain (N=41,677); knee pain (N=77,996); abdominal pain (N=21,285); facial pain (N=4,037); headache (N=39,283); widespread pain (N=6,063). Sample characteristics are further detailed in Supplementary Table S1.

We performed GWAS using REGENIE (v1.0.6.2)^42^ to test associations between the chronic pain types and genetic variants. REGENIE is a GWAS method that implements a logistic mixed model, whereby a genetic relationship between individuals is modelled as a random effect to account for cryptic relatedness. Covariates for the association analysis comprised sex, age, genotyping array and the top 10 principal components derived from genetic data. We used quality control procedures comprising exclusion of variants with: **(i)** minor allele frequency (MAF) < 0.005; **(ii)** imputation quality < 0.6; and **(iii)** deviation from Hardy-Weinberg equilibrium (P-value < 1 × 10^-5^). Subsequently, 11,172,285 single nucleotide polymorphisms (SNPs) remained for analysis. Participant data were excluded if their genotype-derived principal components 1 & 2 were more than six standard deviations from those of the 1000 Genomes European population. Following quality control, up to 441,088 participants remained in the GWAS.

### Genetic Correlations

Genetic correlations were estimated between the regional chronic pain types and a wide range of other complex traits. Cross-trait bivariate linkage disequilibrium (LD) score regression was used — a method that leverages the expected relationship between LD and GWAS association statistics to estimate genetic correlations across traits while accounting for potential sample overlap across studies,^43^ as implemented in CTG-VL.^41^ CTG-VL is a publicly available web platform compiled with >1,490 GWAS summary statistics that is used for running analyses across a comprehensive range of complex traits & disorders (mostly from UK Biobank releases). CTG-VIEW (https://view.genoma.io) is a data aggregator that enables direct queries, visualisations and storage of VL analysis results (and that from other data resources).

A genetic correlation (*rG*) ranges from -1 to 1 and quantifies the genome-wide genetic concordance between a pair of traits. Genetic correlation estimates approaching 1 or -1 indicate (respectively) that a proportion of genetic variants have concordant or divergent effects on both traits. Conversely, a genetic correlation that approaches 0 indicates there is little genetic concordance between the traits or that the genetic predictors of the traits are largely independent. A Benjamini-Hochberg false discovery rate (FDR < 5%) procedure was applied to genetic correlations between complex traits and each chronic pain type GWAS to account for multiple testing.

### Latent Causal Variable Analyses

For trait pairs with non-zero genetic correlation (FDR < 5%), we employed latent causal variable (LCV) analyses to determine if the genetic correlation reflected a causal relationship or horizontal pleiotropy by estimating genetic causal proportion (GCP).^44,45^ The LCV method is robust to horizontal pleiotropy, which is ubiquitous in complex traits. LCV analyses can be conceptualised as assuming a latent ‘causal’ variable that mediates the genetic correlation of two traits. Using mixed fourth moments of the bivariate effect size distribution across all SNPs in both GWAS, LCV estimates the posterior mean for GCP, which indicates the degree to which one trait is correlated with the latent ‘causal’ variable. GCP estimates range from -1 to 1: high positive values (GCP > 0.6) indicate greater partial genetic causality for Trait A (i.e., a chronic pain type) on Trait B (i.e., biological / psychological / social trait); whereas negative values (GCP < -0.6) indicate greater partial genetic causality for Trait B on Trait A. That is, GCP > 0.6 indicates that chronic pain likely has a causal effect on another particular trait, whereas GCP < -0.6 indicates a particular trait likely has a causal effect on chronic pain. Benjamini-Hochberg false discovery rate (FDR < 5%) was applied across GCPs for complex traits with each chronic pain type GWAS.

## Results

### Genetic Correlations & Latent Causal Variable Analyses

For each of the eight chronic pain types, details of significant genetic correlations with putative causal relationships are summarised in Figures 2–8 and Supplementary Figures S1 & S2, while *rG* and GCP data are reported in Supplementary Tables S2–S9. Complete genetic correlation and LCV analysis results for all 1,492 traits are presented in Supplementary Tables S10–S17.

**Figure 2:**
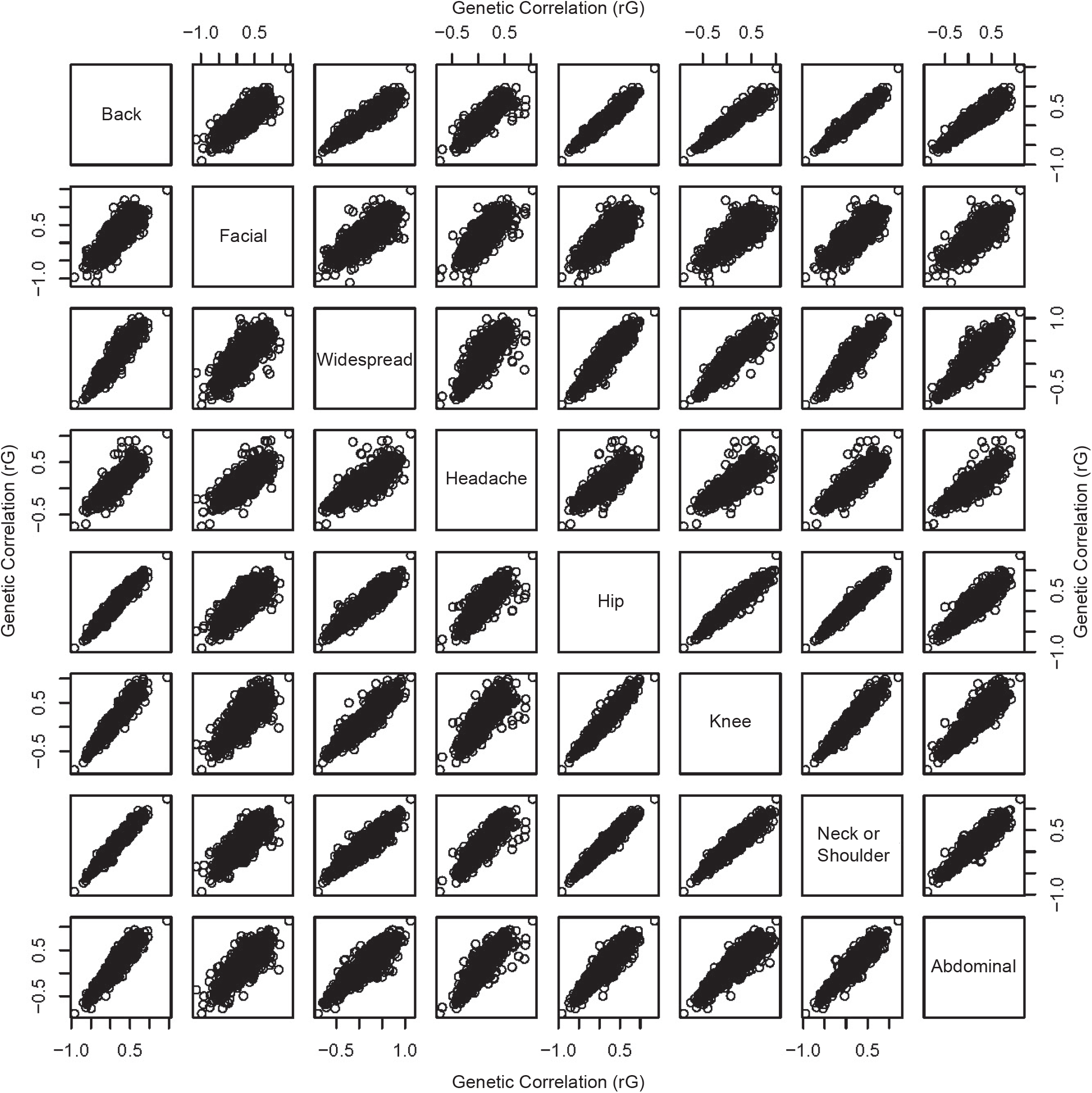
Scatterplot matrix showing the genetic correlations (*rG*) between chronic pain and 1,492 complex traits across the eight regional pain types. The genetic correlations were broadly consistent across these types, which indicates the genetic variants specific to each of the 1,492 complex traits had consistent correlations (positive or negative) with the genetic variants associated with each of the chronic pain types. This pattern of results points toward a shared genetic basis across these chronic pain types, which may partly account for both an increased risk of developing chronic pain (more generally) and its commonly comorbid biopsychosocial traits.

**Figure 3:**
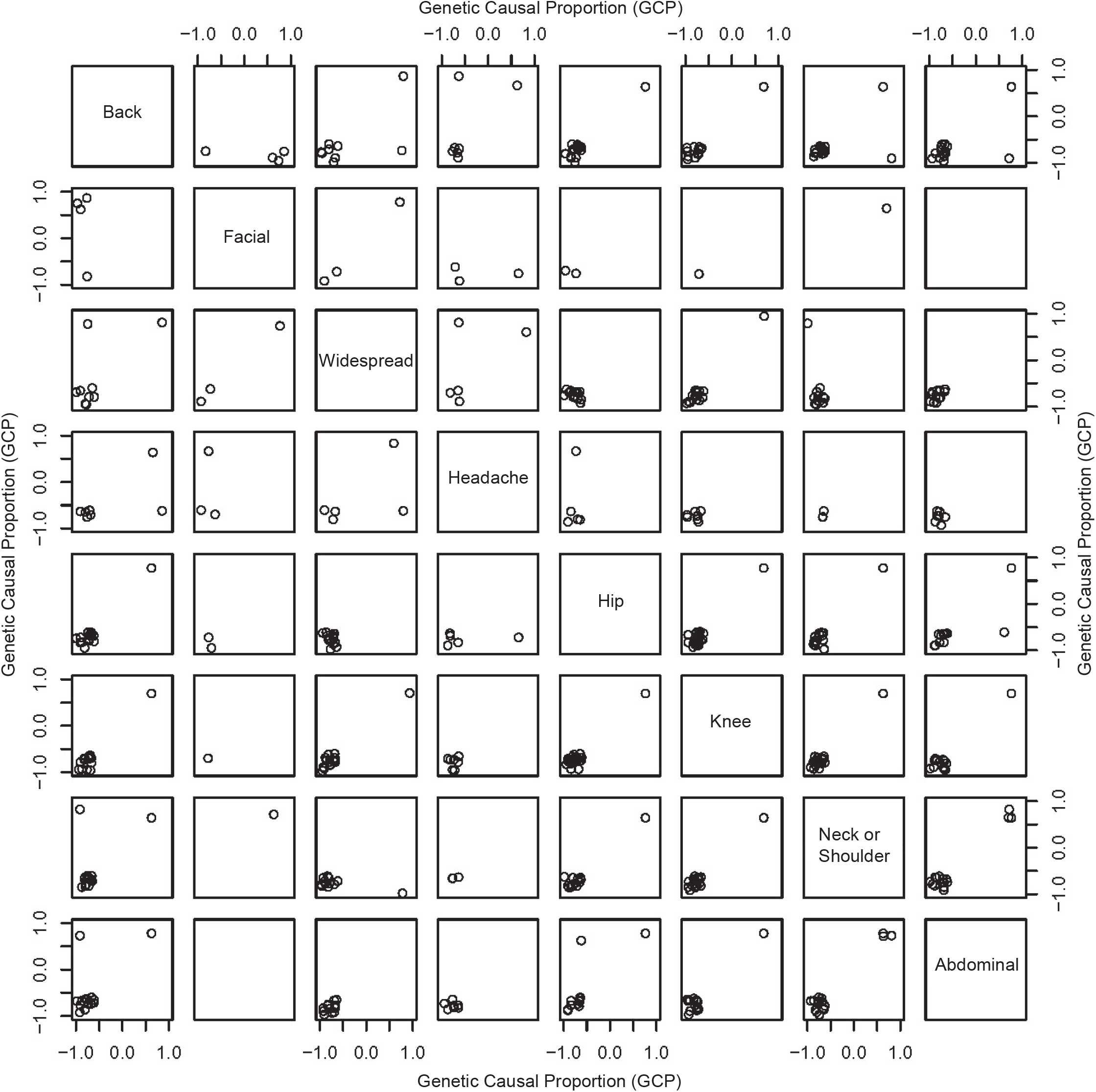
Scatterplot matrix showing 570 significant genetic causal proportions (GCPs) between chronic pain and 1,492 complex traits across the eight regional pain types wherein the LCV analysis method took into account the overlapping samples. There were eight instances where the direction of causal relationships (denoted by GCP > 0.6 or < - 0.6) differed between the pain types as detailed in Supplementary Table S18. These included psychological traits (e.g., concerning mania/irritability), family history/social factors, musculoskeletal pathology & other clinical traits. Such differences in the traits’ casual directions across pain types may reflect broader multi-directional dynamic relationships (e.g., mania/irritability both impacting & being affected by chronic pain) or may be more specific to regional pain types (e.g., arthrosis contributing to hip pain as joint pathology cf. abdominal pain contributing to arthrosis indirectly, such as via physical inactivity or another pathway). The otherwise broadly consistent direction of causal relationships with complex traits across these common chronic pain types supports a shared biopsychosocial genetic basis.

**Figure 4:**
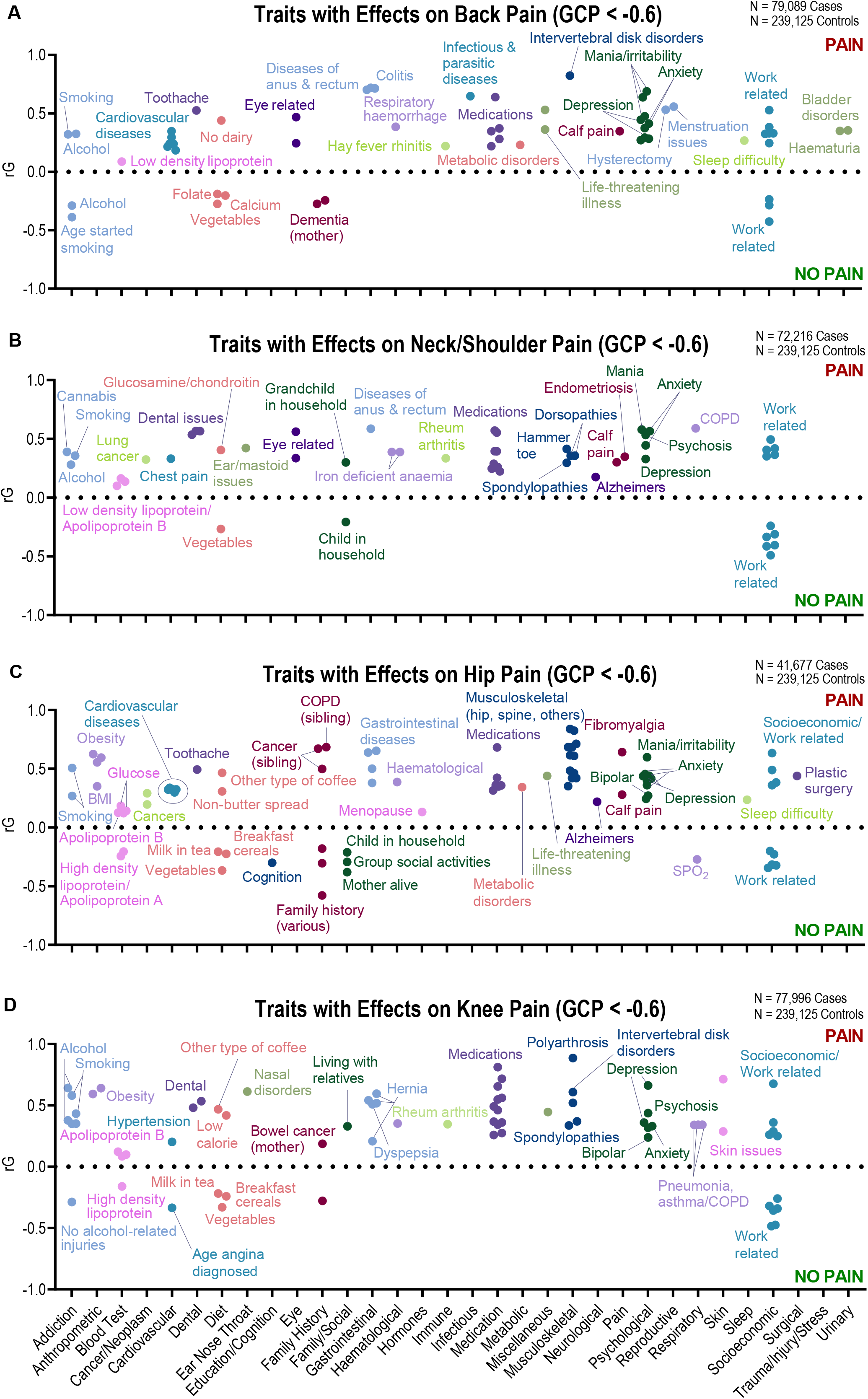
Genetic correlations (*rG*) for traits with significant genetic causal proportion (GCP) estimates indicative of a causal effect upon regional chronic pain types (GCP < - 0.6, 5% false discovery rate) are shown across the range of complex trait categories (x-axis). Data are presented for (**A**) chronic back pain, (**B**) neck/shoulder pain, (**C**) hip pain and (**D**) knee pain. A broad range of traits demonstrate evidence of genetic causal effects contributing to an increased risk of chronic pain — notably, psychological traits (e.g., depression on back, neck/shoulder, hip & knee pain), socioeconomic factors (e.g., work-related traits on back, neck/shoulder, hip & knee pain), musculoskeletal pathology (e.g., spondylopathies on neck/shoulder pain), medication use (e.g., dihydrocodeine on back pain) and addiction (e.g., smoking on back, neck/shoulder, hip & knee pain). Other traits had an apparent protective effect against these chronic pain types such as vegetable & dairy consumption and certain work-associated factors (e.g., software programmer/engineer and reduced risk of neck/shoulder pain — Supplementary Table S3 & Supplementary Results 2). BMI: body mass index; COPD: chronic obstructive pulmonary disease.

**Figure 5:**
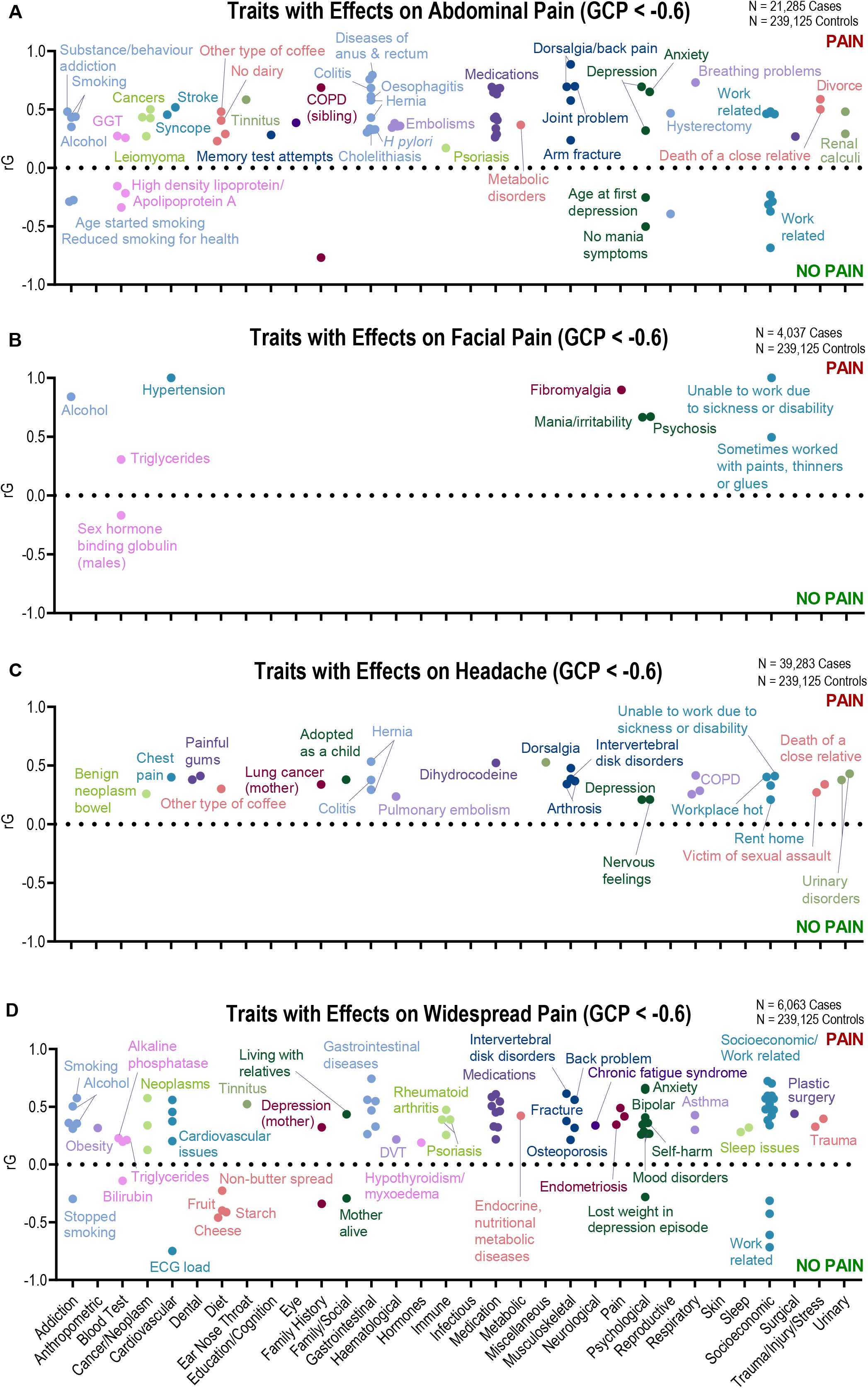
Genetic correlations (*rG*) for traits with significant genetic causal proportion (GCP) estimates indicative of a causal effect upon regional chronic pain types (GCP < - 0.6, 5% false discovery rate) are shown across the range of complex trait categories (x-axis). Data are presented for (**A**) chronic abdominal pain, (**B**) facial pain, (**C**) headaches and (**D**) widespread pain. A broad range of traits demonstrate evidence of genetic causal effects on these pain types, including psychological (e.g., depression, trauma/stress, mania and/or anxiety on headaches, abdominal, facial & widespread pain), gastrointestinal (e.g., colitis on headaches & abdominal pain) and socioeconomic factors (e.g., work-related traits on headaches, abdominal, facial & widespread pain). Other traits had an apparent protective effect against certain chronic pain types such as dairy & cheese consumption (abdominal & widespread pain, respectively) and working as a teacher (abdominal & widespread pain). COPD: chronic obstructive pulmonary disease; DVT: deep vein thrombosis; ECG: electrocardiogram; GGT: gamma-glutamyl transferase.

**Figure 6:**
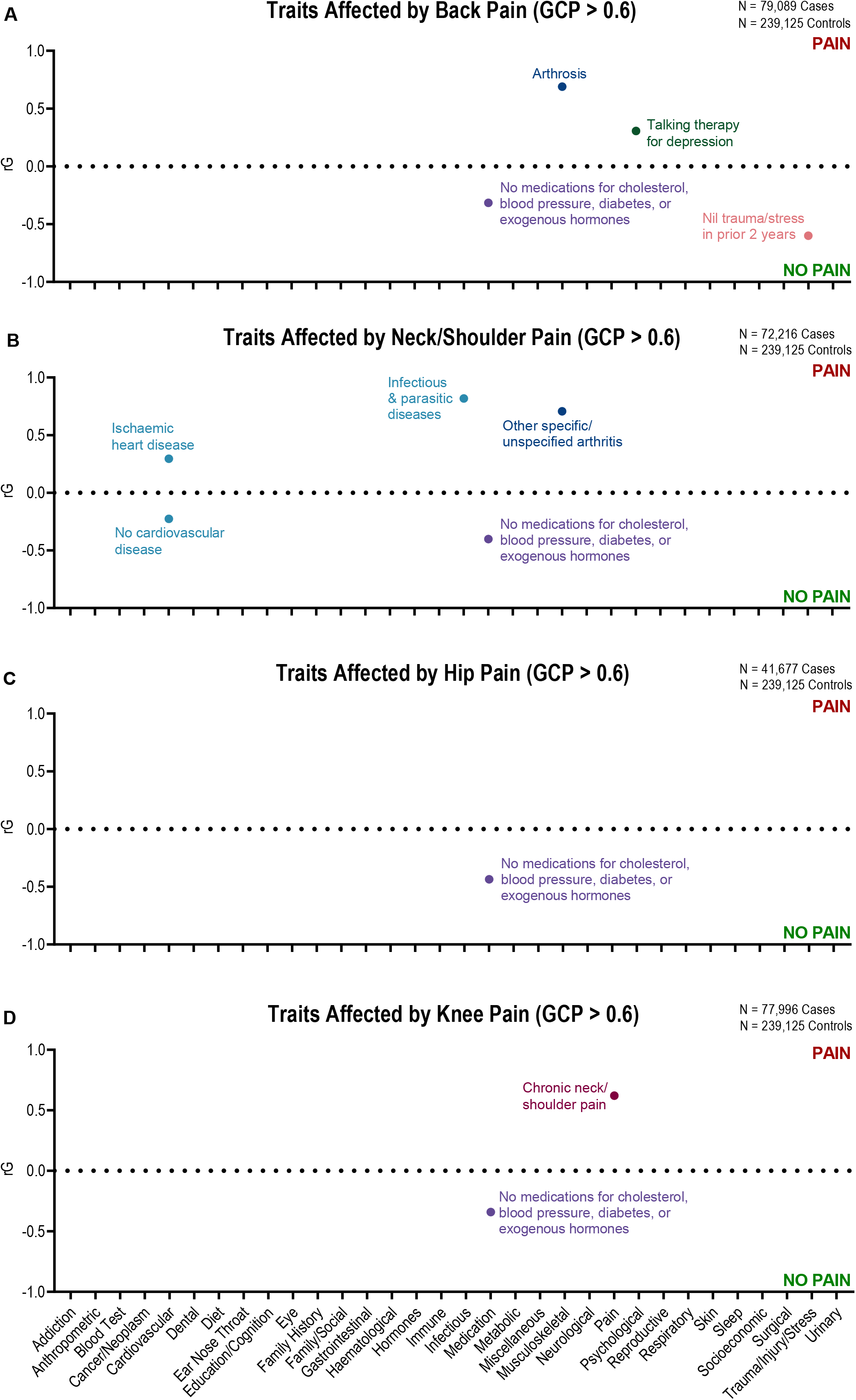
Genetic correlations (*rG*) for traits with significant genetic causal proportion (GCP) estimates indicative of being affected by chronic pain (GCP > 0.6, 5% false discovery rate) are shown across the range of complex trait categories (x-axis). Data are presented for (**A**) chronic back pain, (**B**) neck/shoulder pain, (**C**) hip pain and (**D**) knee pain. While there are fewer instances of these regional chronic pain affecting other traits than *vice versa* (Fig. 4), the results suggest these chronic pain types have genetic causal effects contributing to increased medication use (i.e., for cholesterol, blood pressure, diabetes or exogenous hormones) and an increased risk of cardiovascular disease (i.e., neck/shoulder pain on ischemic heart disease) along with other disorders (e.g., back pain on depression).

**Figure 7:**
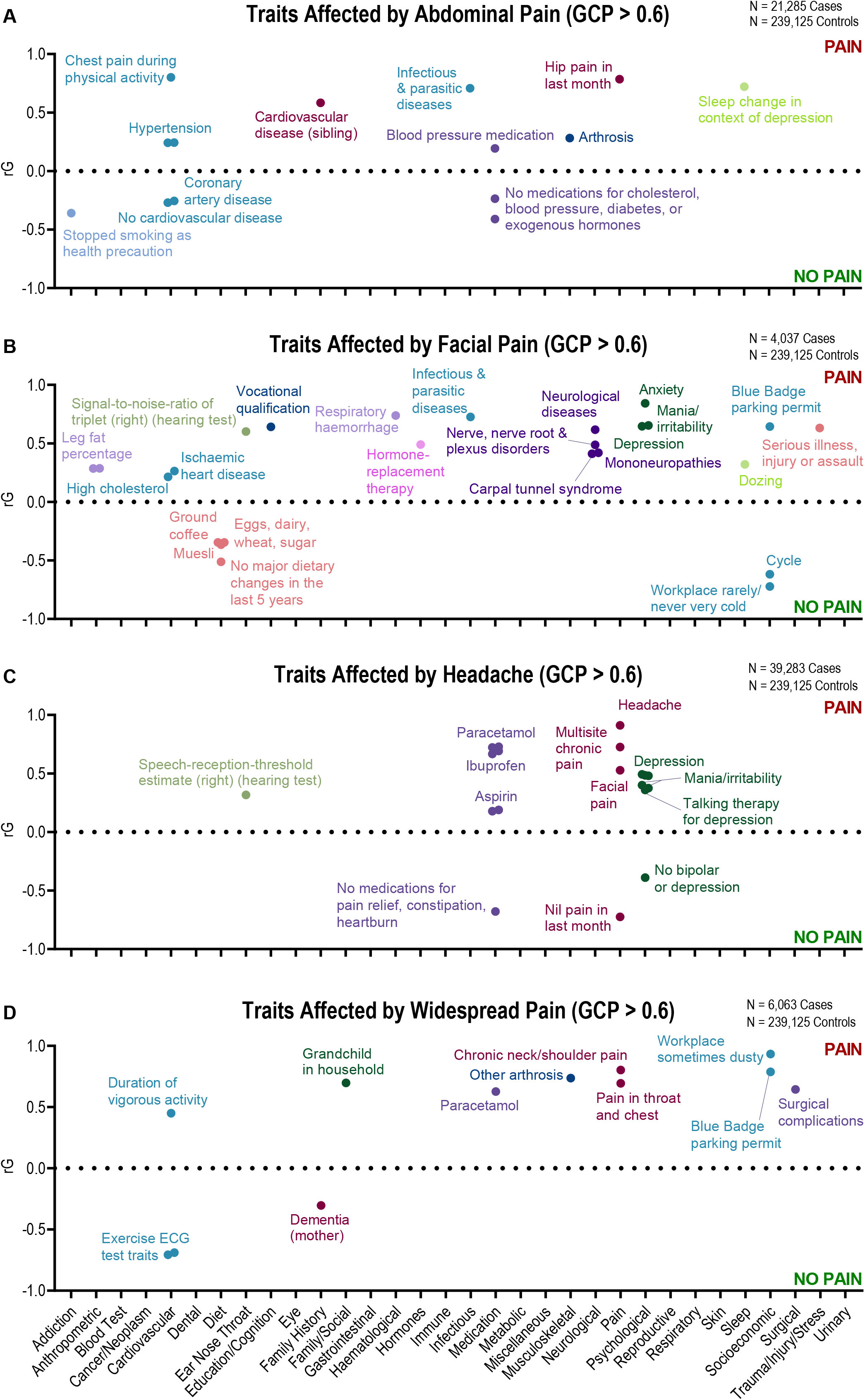
Genetic correlations (*rG*) for traits with significant genetic causal proportion (GCP) estimates indicative of being affected by chronic pain (GCP > 0.6, 5% false discovery rate) are shown across the range of complex trait categories (x-axis). Data are presented for (**A**) chronic abdominal pain, (**B**) facial pain, (**C**) headaches and (**D**) widespread pain. Notably, facial pain showed effects on a wide range of traits including dietary patterns (e.g., reduced muesli, ground coffee), psychological factors (e.g., increased depression, anxiety) and an increased risk of neurological conditions (e.g., mononeuropathies, carpal tunnel syndrome), whereas abdominal pain particularly contributed to an increased risk of cardiovascular disease (e.g., hypertension). Chronic headache had genetic effects contributing to increased analgesic use (i.e., paracetamol, ibuprofen, aspirin) & psychological traits (e.g., depression). Widespread pain affected a range of traits such as an increased risk of surgical complications, analgesic use and disability parking permit use. ECG: electrocardiogram.

**Figure 8:**
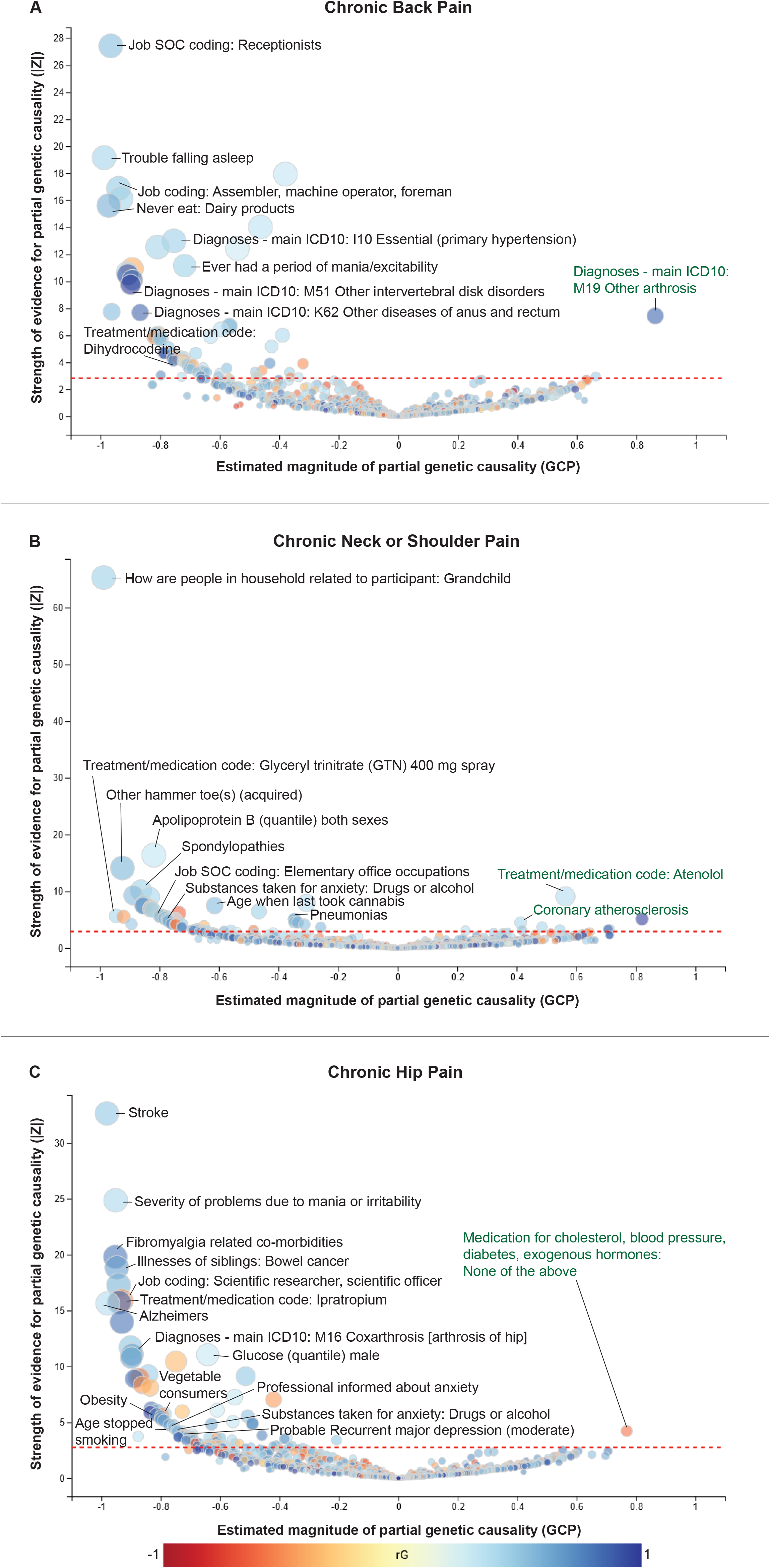
Causal architecture plots presenting the relationships of various traits with (**A**) chronic back pain, (**B**) neck/shoulder pain and (**C**) hip pain. Each dot represents a genetic correlation (*rG*). The colour of a dot indicates whether the genetic correlation between them is positive (red) or negative (blue). The x-axis denotes the genetic causal proportion (GCP) estimate and the y-axis denotes the GCP absolute Z-score (i.e., statistical significance). The red dashed line shows the threshold for statistical significance (5% false discovery rate). Traits that have a genetic causal effect on chronic pain are on the left side of the plot (with respect to 0 on the x-axis, GCP < -0.6; black text), while traits causally affected by chronic pain are on the right side (GCP > 0.6; green text). Selected traits are labelled for illustrative purposes — spanning clinical, biological, psychological & socioeconomic measures. Full details of all significant genetic causal relationships are provided in Supplementary Tables S2, S3 & S4. Causal architecture plots for the other regional chronic pain types are presented in Supplementary Figures S1 & S2.

From 11,932 trait pairs, we identified 5,942 significant genetic correlations (which corroborate many reported phenotypic associations from previous clinical studies), of which 570 trait pairs showed evidence of a causal association (|GCP| > 0.6; 5% false discovery rate). Of these, 488 traits had genetic casual effects on chronic pain, while the remaining 82 traits were affected by pain.

Comparisons of *rG* and significant GCP results across the eight chronic pain types are presented in Figures 2 & 3. Overall, the direction of the genetic correlations between the complex traits and chronic pain were broadly consistent across all eight types. There were eight instances where the direction of genetic causal relationships (denoted by GCP > 0.6 or < -0.6) differed between the chronic pain types (Supplementary Table S18). Additional *rG* estimates for the eight chronic pain types are presented in Supplementary Table S19, including significant genetic correlations between each type, whereby the analysis method accounted for the overlapping samples.

### Key Results Across Chronic Pain Types

Significant *rG* (5% FDR) and GCP estimates (< -0.6; 5% FDR) were observed for musculoskeletal pathologies, which showed genetic causal effects on chronic back, neck/shoulder, hip, knee, abdominal, headaches and widespread pain, including local sites (e.g., coxarthrosis of the hip & chronic hip pain *rG* [S.E.] = 0.42 [0.06]; GCP [S.E.] = -0.90 [0.08]) as well as remote sites (e.g., other intervertebral disc disorders & chronic knee pain *rG* [S.E.] = 0.61 [0.12], GCP [S.E.] = -0.79 [0.16]) (Figures 4–8, Supplementary Figures S1 & S2, Supplementary Tables S2–S9). Cardiovascular disease-associated traits affected each of the chronic pain types (e.g., heart failure contributes to back pain [*rG* (S.E.) = 0.22 (0.09); GCP (S.E.) = -0.73 (0.16)] and hip pain [*rG* (S.E.) = 0.30 (0.11); GCP (S.E.) = -0.62 (0.24)]), while certain chronic pain types (i.e., neck/shoulder, abdominal, facial & widespread pain) also affected cardiovascular traits such as ischaemic heart disease (neck/shoulder pain *rG* [S.E.] = 0.30 [0.04], GCP [S.E.] = 0.71 [0.22]; facial pain *rG* [S.E.] = 0.26 [0.09], GCP [S.E.] = 0.64 [0.22]). A number of chronic pain types (i.e., back, neck/shoulder, hip, knee, abdominal & headaches) contributed to medication use for cholesterol, blood pressure, diabetes or exogenous hormones. Chronic headaches and widespread pain also impacted the use of analgesics (i.e., paracetamol, ibuprofen, aspirin; and paracetamol, respectively), while the use of various medications (e.g., analgesics, cardiovascular drugs) affected chronic back, neck/shoulder, hip, knee, abdominal, headache and widespread pain.

Psychological traits and psychiatric diagnoses also had genetic causal effects on chronic pain types. Notably: **(i)** depression and anxiety-associated traits increased the risk of chronic back, neck/shoulder, hip, knee, abdominal, headache and widespread pain; **(ii)** mania-associated traits increased the risk of chronic back, neck/shoulder, hip, knee, facial and widespread pain; and **(iii)** substance abuse/addiction-associated traits increased the risk of chronic back, neck/shoulder, hip, knee, abdominal, facial and widespread pain. Traits concerning bipolar disorder and psychosis respectively increased the risk of hip, knee and widespread pain; and of neck/shoulder, knee and facial pain. Chronic back pain, facial pain and headaches had genetic causal effects upon traits concerning depression, anxiety and mania (significant *rG* [5% FDR] and GCP > 0.6 [5% FDR]). Traits specific to occupations demonstrated genetic causal contributions to back, neck/shoulder, hip, knee, abdominal and widespread pain (e.g., receptionist & chronic back pain *rG* [S.E.] = 0.38 [0.17]; GCP [S.E.] = -0.97 [0.04]), while traits relating to work environment increased the risk of all eight chronic pain types (e.g., *‘Workplace had a lot of diesel exhaust: Often’* and chronic widespread pain *rG* [S.E.] = 0.60 [0.15]; GCP [S.E.] = -0.81 [0.15]).

### Region Specific Summaries

Additional results on the key findings for each pain type are presented in the Supplementary Results.

## Discussion

The multifactorial nature of chronic pain conditions means that assessing their aetiological relationships is challenging due to the large sample sizes and long follow-up timeframes required for multivariate longitudinal analyses. Large-scale genetic datasets adequately powered for GWAS, along with statistical genetics methods developed for investigating causality (e.g., LCV, Mendelian randomization [MR]), present a way forward. The primary findings of this data-driven study provide quantitative genetic evidence for a range of shared biopsychosocial factors demonstrating causal effects upon eight common types of chronic pain. Certain clinical factors (e.g., musculoskeletal pathology, medical comorbidities), psychological/psychiatric traits (e.g., depression, trauma, anxiety, mania) and socioeconomic status (e.g., occupation) increased the risk of chronic pain types. The results also indicated genetic causal effects of chronic pain upon other traits such as increased medication use and increased risk of cardiovascular disease & depression. The current study has characterised the multidimensional casual traits and substantially narrowed down the range of genetically supported risk and protective factors for chronic pain. This work lays the foundation for further translational studies to identify the underlying causal genetic variants and pathophysiological pathways for developing novel diagnostic & therapeutic technologies and strategies.

Overall, the directions of genetic correlations were consistent across an array of biopsychosocial traits and common chronic pain types. The vast majority of significant genetic causal directions between them were also consistent, while only a small minority differed across the chronic pain types — which suggests an underlying universal genetic core that is shared with these complex traits. Significant genetic correlations found between each of the eight chronic pain types provide additional evidence of a shared genetic signature, which builds upon recent studies^5,46^ and earlier proposals^47^ pursuing this empirical demonstration. The biopsychosocial traits identified in the current study can also serve as candidates for patient classification, risk stratification and pain phenotyping in the clinical management of chronic pain conditions.^7^ Furthermore, based on the precedent for genetically supported therapeutic targets,^3,4^ the identified traits with genetic causal effects are potential targets in the clinical management of chronic pain as our results provide corroborating genetic evidence for current treatment & prevention strategies (e.g., weight-loss in knee osteoarthritis^48^ & targeting post-traumatic stress after injury^49,50^, respectively).

The results also indicate corroborating genetic evidence that musculoskeletal pathologies (e.g., intervertebral disc disorders in back pain, spondylopathies in neck/shoulder pain, coxarthrosis in hip pain) and visceral pathologies (e.g., renal & ureteric calculi) contribute to chronic pain. We found genetic causal effects of painful pathologies at remote sites upon regional chronic pain types (e.g., shoulder impingement on chronic hip pain), which is consistent with a shared genetic signature and associated predisposition to chronic pain (i.e., an underlying genetic predisposition to musculoskeletal shoulder pathology also contributes to increased risk of chronic hip pain).

Not surprisingly, the results indicate that certain chronic pain types (i.e., headaches & widespread pain) contribute to use of analgesic medications (i.e., paracetamol, ibuprofen, aspirin; and paracetamol, respectively). Interestingly, various drugs used in the treatment of pain (e.g., opioids, non-steroidal anti-inflammatory drugs, antidepressants^27^), cardiovascular disease (e.g., statins, beta-blockers) and gastro-oesophageal reflux showed an effect of increased risk for various chronic pain types. While some of these may reflect a genetic correlation between pain and problematic opioid use,^51^ the effect of medication use on increasing the risk of chronic pain may also be attributable to an indirect causal effect. For example, an unobserved trait (e.g., symptom/disease), which increases the likelihood of both medication use and a chronic pain type, could create an apparent casual effect between that medication and the pain type. That is, medication use (e.g., analgesics) could simply serve as a proxy for conditions that cause pain or that are commonly co-morbid with chronic pain. For instance, antidepressants may reflect the relationship between chronic pain and depression,^27^ or conversely, the use of antidepressants to treat chronic pain conditions.^52,53^

Our findings indicate that while pain conditions and psychological traits (e.g., depression) are frequently co-morbid at a phenotypic level, the pattern of causal relationships between them are heterogenous across the various chronic pain types. For instance, we observed genetic evidence of causal relationships across both directions between depression-associated traits and chronic headaches & back pain (i.e., a proportion of relationships are pain → depression and others are depression → pain [Supplementary Tables S2, S8]). In contrast, depression-associated traits exclusively showed effects on chronic neck/shoulder, hip, knee, abdominal and widespread pain (i.e., depression → pain only [Supplementary Tables S3–6, S9]), consistent with evidence for (modest) effects of treatments targeting depression upon pain & disability in such conditions (e.g., serotonin-noradrenaline reuptake inhibitors for knee osteoarthritis^53^). Conversely, chronic facial pain increased the risk of seeking care for depression (i.e., pain → depression only [Supplementary Table S7]). These findings are broadly consistent with other genetic studies concerning depression and chronic pain, which have shown genetic causal relationships between chronic pain and depression, spanning both depression → pain pathways^54–58^ and pain → depression pathways.^20,52,55,59^ Taken together, our results and previous findings^19,20,27,54,55^ raise further intriguing hypotheses, such as that either or both causal directions may exist on a patient sub-group basis.^52^ Our LCV results also implicate a history of trauma or stress — including death of a close relative in last two years; being adopted as a child; sexual assault; marital separation; and military service — as genetic causal factors for various chronic pain types. Phenotypic studies indicate that trauma^60,61^ or post-traumatic stress^62^ are risk factors for development of chronic pain.^63^ Our LCV results provide convergent evidence for targeting post-traumatic stress symptoms in treatment of pain, such as recently reported in a randomised controlled trial for whiplash-associated disorder,^49^ which showed reductions in pain and disability through improvements in stress, depression and post-traumatic stress.^50^ In addition to depression and trauma, traits concerning anxiety, psychosis, bipolar disorder and mania showed genetic causal effects on several chronic pain types.

Socioeconomic factors also demonstrated genetic causal effects on a range of chronic pain types. Broadly, occupational traits associated with lower socioeconomic status (e.g., elementary office worker) contributed to increased chronic pain risk, whereas those with higher incomes (e.g., software engineer) were protective against chronic pain. Socioeconomic status may affect pain risk^33^ through a range of potential pathways, including increased likelihood of: **(i)** lower educational attainment (associated with use of less effective pain coping strategies^64^); **(ii)** various negative health behaviours (e.g., low physical activity & smoking^65^); **(iii)** early life adverse events^66^; and **(iv)** impaired access to healthcare.^67^ Accordingly, interventions and policies targeting socioeconomic inequality may have beneficial downstream effects on the broader community burden of chronic pain.

Various medical conditions such as cardiovascular, gastrointestinal & respiratory diagnoses demonstrated genetic correlations and genetic causal relationships with chronic pain types. Notably, cardiovascular-related traits showed heterogeneous associations. For example, heart failure contributed to chronic back and hip pain, whereas neck/shoulder and facial pain increased the risk of ischaemic heart disease. Numerous cardiovascular risk factors showed genetic causal contributions to chronic pain such as obesity (i.e., hip, knee & widespread pain), low-density lipoprotein/Apolipoprotein B (i.e., back, neck/shoulder, hip & knee pain) and triglycerides (i.e., facial & widespread pain). These findings are consistent with phenotypic relationships between chronic pain and cardiovascular diagnoses/mortality, which may reflect other associations between chronic pain and cardiac risk factors (e.g., smoking).^37^

A limitation of the current study is the minimal clinical details used to define regional chronic pain (i.e., a self-report questionnaire regarding presence of pain for >3 months at a body site). However, this minimal phenotyping approach enables acquisition of large-scale data with sufficient power for GWAS and post-GWAS analyses of causal relationships. As the chronic pain and complex trait GWAS data were mostly derived from the UK Biobank, we employed LCV analyses rather than a more traditional approach (e.g., MR) because overlapping samples in the latter can bias estimates towards the confounded associations.^68^ Although the LCV method is not able to detect bidirectional relationships between two individual traits (e.g., chronic pain → Trait A and Trait A → chronic pain), its current application in comprehensive phenome-wide analyses — particularly for complex disorders such as chronic pain — is an efficient & effective strategy upon which to follow up with traditional approaches (e.g., MR, genomic structural equation modelling^69^) targeting the genetic causal traits delineated in the current study.

Further work is required including: **(i)** genetic epidemiology studies collecting data on more specific chronic pain diagnoses, clinical traits (e.g., pro- & anti-nociceptive phenotypes, quantitative sensory testing,^70–73^ medication/treatment responses^52,74^) and epigenetic factors^75^ (e.g., early life stress, physical activity); **(ii)** replication in other large genotyped datasets,^5,19^ particularly population samples with non-European ancestry; and **(iii)** incorporating putative transdiagnostic subtypes and genetic risk stratification of placebo/control groups in clinical trials of chronic pain interventions.^3,4,27,52,76^

Our LCV analysis of >1,400 biopsychosocial complex traits has leveraged the underlying genetic variant data to provide a comprehensive characterisation of risk & protective factors across common regional chronic pain conditions, and in turn, testable hypotheses for further targeted interrogation. It has also provided corroborating genetic evidence for current treatment approaches (e.g., targeting psychological traits), which lays the foundation for further translational studies into developing more effective multidisciplinary care frameworks and enhancing patient outcomes.

## Data Statement

Complex trait GWAS summary statistics are publicly available. Links to reference and download each of these are available in CTG-VL (https://vl.genoma.io) and CTG-VIEW (https://view.genoma.io). Complete GWAS summary statistics and results from downstream analyses for all the chronic pain types reported in the current study (Supplementary Table S1) are available at CTG-VIEW.

## Supporting information

Supplementary Results

STREGA

## Supplementary Tables

**Supplementary Table S1:** Distribution of chronic pain across body sites, sex and prevalence in the sample.

**Supplementary Table S2:** Genetic correlation (*rG*) and genetic causal proportion (GCP) results for traits with significant causal relationships with chronic back pain.

**Supplementary Table S3:** Genetic correlation (*rG*) and genetic causal proportion (GCP) results for traits with significant causal relationships with chronic neck/shoulder pain.

**Supplementary Table S4:** Genetic correlation (*rG*) and genetic causal proportion (GCP) results for traits with significant causal relationships with chronic hip pain.

**Supplementary Table S5:** Genetic correlation (*rG*) and genetic causal proportion (GCP) results for traits with significant causal relationships with chronic knee pain.

**Supplementary Table S6:** Genetic correlation (*rG*) and genetic causal proportion (GCP) results for traits with significant causal relationships with chronic abdominal pain.

**Supplementary Table S7:** Genetic correlation (*rG*) and genetic causal proportion (GCP) results for traits with significant causal relationships with chronic facial pain.

**Supplementary Table S8:** Genetic correlation (*rG*) and genetic causal proportion (GCP) results for traits with significant causal relationships with chronic headaches.

**Supplementary Table S9:** Genetic correlation (*rG*) and genetic causal proportion (GCP) results for traits with significant causal relationships with chronic widespread pain.

**Supplementary Table S10:** Genetic correlation (*rG*) and genetic causal proportion (GCP) results for 1,492 traits with chronic back pain.

**Supplementary Table S11:** Genetic correlation (*rG*) and genetic causal proportion (GCP) results for 1,492 traits with chronic neck/shoulder pain.

**Supplementary Table S12:** Genetic correlation (*rG*) and genetic causal proportion (GCP) results for 1,492 traits with chronic hip pain.

**Supplementary Table S13:** Genetic correlation (*rG*) and genetic causal proportion (GCP) results for 1,492 traits with chronic knee pain.

**Supplementary Table S14:** Genetic correlation (*rG*) and genetic causal proportion (GCP) results for 1,492 traits with chronic abdominal pain.

**Supplementary Table S15:** Genetic correlation (*rG*) and genetic causal proportion (GCP) results for 1,492 traits with chronic facial pain.

**Supplementary Table S16:** Genetic correlation (*rG*) and genetic causal proportion (GCP) results for 1,492 traits with chronic headache.

**Supplementary Table S17:** Genetic correlation (*rG*) and genetic causal proportion (GCP) results for 1,492 traits with chronic widespread pain.

**Supplementary Table S18:** Traits demonstrating contrary directions of causal relationships between chronic pain types.

**Supplementary Table S19:** Genetic correlation (*rG*) results between chronic pain types.

## Supplementary Figures

**Supplementary Figure S1:** Causal architecture plots presenting the relationships of various traits with (**A**) chronic knee pain, (**B**) widespread pain and (**C**) headaches. Each dot is a data point representing a genetic correlation (*rG*). The colour of a dot indicates whether the genetic correlation between them is positive (red) or negative (blue). The x-axis denotes the genetic causal proportion (GCP) estimate and the y-axis denotes the GCP absolute Z-score (i.e., statistical significance). The red dashed line shows the threshold for statistical significance (5% false discovery rate). Traits that have a genetic causal effect on chronic pain are on the left side of the plot (with respect to 0 on the x-axis; GCP < -0.6, black text), while traits causally affected by chronic pain are on the right side (GCP > 0.6, green text). Selected traits are labelled for illustrative purposes — spanning clinical, biological, psychological & socioeconomic measures. Full details of all significant genetic causal relationships are provided in Supplementary Tables S5, S8 & S9.

**Supplementary Figure S2:** Causal architecture plots presenting the relationships of various traits with (**A**) chronic abdominal pain and (**B**) facial pain. Each dot is a data point representing a genetic correlation (*rG*). The colour of a dot indicates whether the genetic correlation between them is positive (red) or negative (blue). The x-axis denotes the genetic causal proportion (GCP) estimate and the y-axis denotes the GCP absolute Z-score (i.e., statistical significance). The red dashed line shows the threshold for statistical significance (5% false discovery rate). Traits that have a genetic causal effect on chronic pain are on the left side of the plot (with respect to 0 on the x-axis; GCP < -0.6, black text), while traits causally affected by chronic pain are on the right side (GCP > 0.6, green text). Selected traits are labelled for illustrative purposes. Full details of all significant genetic causal relationships are provided in Supplementary Tables S6 & S7.

