## Supplementary Results for "A shared genetic signature for common chronic pain conditions and its impact on biopsychosocial traits"

### Additional Reporting of Results

*Chronic Back Pain*
We found significant genetic correlations between chronic back pain and 841/1492 traits (FDR < 5%) (Supplementary Table S10). Sixty (60) traits had genetic correlations with putative casual effects on chronic back pain risk (GCP < -0.6), while four other traits were causally affected by chronic back pain (GCP > 0.6) (Figures 4, 6 & 8, Supplementary Table S2). A particular finding to note — in addition to the key results reported in the main text — was that dietary factors (e.g., vegetable consumption, dairy consumption, calcium, folate) had a protective effect on the risk of chronic back pain, whereas 'trouble falling asleep' contributed to increased risk of chronic back pain (*rG* [S.E.] = 0.27 [0.07]; GCP [S.E.] = -0.99 [0.05]).

*Chronic Neck/Shoulder Pain*
Significant genetic correlations were found between chronic neck/shoulder pain and 863/1492 traits (FDR < 5%) (Supplementary Table S11). Fifty-five (55) traits had genetic correlations with putative casual effects on chronic neck/shoulder pain risk, while five other traits were causally affected by chronic neck/shoulder pain (Figures 4, 6 & 8, Supplementary Table S3). It is worth noting that in addition to the key results reported in the main text, comorbid medical conditions showed genetic causal effects upon neck/shoulder pain, including chronic obstructive pulmonary disease (*rG* [S.E.] = 0.59 [0.11]; GCP [S.E.] = -0.66 [0.23]) and Alzheimer's disease (*rG* [S.E.] = 0.17 [0.08]; GCP [S.E.] = -0.63 [0.24]), along with (not) consuming vegetables (*rG* [S.E.] = -0.27 [0.11]; GCP [S.E.] = -0.83 [0.12]) and various dental problems.

*Chronic Hip Pain*
We found significant genetic correlations between chronic hip pain and 849/1492 traits (FDR < 5%) (Supplementary Table S12). Eighty-six (86) traits had genetic correlations with putative casual effects on chronic hip pain risk, while one other trait was causally affected by chronic hip pain (Figures 4, 6 & 8, Supplementary Table S4). Notably, a range of traits related to obesity (including BMI) also showed genetic causal effects on chronic hip pain.

*Chronic Knee Pain*
Significant genetic correlations were found between chronic knee pain and 882/1492 traits (FDR < 5%) (Supplementary Table S13). Seventy-three (73) traits had genetic correlations with putative casual effects on chronic knee pain, while two other traits were causally affected by chronic knee pain (Figures 4 & 6, Supplementary Figure S1, Supplementary Table S5). Notably, traits related to obesity and abdominal/gastrointestinal conditions (e.g., dyspepsia & hernias) contributed to an increased risk of chronic knee pain.

*Chronic Abdominal Pain*
We found significant genetic correlations between chronic abdominal pain and 743/1492 traits (FDR < 5%) (Supplementary Table S14). Eighty-one (81) traits had genetic correlations with putative casual effects on chronic abdominal pain risk, while 14 other traits were causally affected by chronic abdominal pain (Figures 5 & 7, Supplementary Figure S2, Supplementary Table S6). A wide range of pathologies & other clinical factors appeared to influence chronic abdominal pain such as hernias, cancers (lung & uterus), renal/ureteral calculi, haematuria, cholelithiasis, ulcerative colitis, Crohn's disease, *Helicobacter pylori* and liver function tests.

*Chronic Facial Pain*
Significant genetic correlations were found between chronic facial pain and 401/1492 traits (FDR < 5%) (Supplementary Table S15). Nine traits had genetic correlations with putative casual effects on chronic facial pain risk, while 25 other traits were causally affected by chronic facial pain (Figures 5 & 7, Supplementary Figure S2, Supplementary Table S7). Notably, chronic facial pain showed mixed relationships with psychological traits. For example, it contributed to depression, anxiety & mania, whereas restlessness (as a manifestation of mania or irritability) (*rG* [S.E.] = 0.67 [0.12]; GCP [S.E.] = -0.76 [0.17]) and hearing an un-real voice (*rG* [S.E.] = 0.67 [0.24]; GCP [S.E.] = -0.67 [0.26]) both showed genetic causal effects on chronic facial pain.

*Chronic Headaches*
We found significant genetic correlations between chronic headaches and 516/1492 traits (FDR < 5%) (Supplementary Table S16). Thirty (30) traits had genetic correlations with putative casual effects on

chronic headache risk, while 19 other traits were causally affected by chronic headaches (Figures 5 & 7, Supplementary Figure S1, Supplementary Table S8). Notably, traits concerning bereavement (e.g., death of a close relative in last 2 years *rG* [S.E.] = 0.34 [0.08]; GCP [S.E.] = -0.74 [0.21]), being adopted as a child (*rG* [S.E.] = 0.38 [0.13]; GCP [S.E.] = -0.98 [0.05]) and sexual assault (*rG* [S.E.] = 0.27 [0.06]; GCP [S.E.] = -0.72 [0.19]) led to increased risk of chronic headaches.

*Chronic Widespread Pain*

Significant genetic correlations were found between chronic widespread pain and 847/1492 traits (FDR < 5%) (Supplementary Table S17). Ninety-four (94) traits had genetic correlations with putative casual effects on chronic widespread pain risk, while 12 other traits were causally affected by chronic widespread pain (Figures 5 & 7, Supplementary Figure S1, Supplementary Table S9). Notably, (not) consuming fruit (*rG* [S.E.] = -0.40 [0.11]; GCP [S.E.] = -0.80 [0.14]), cheese (*rG* [S.E.] = -0.46 [0.15]; GCP [S.E.] = -0.78 [0.16]) and starch (*rG* [S.E.] = -0.41 [0.17]; GCP [S.E.] = -0.74 [0.18]) increased the risk of chronic widespread pain, as did 'trouble falling asleep' (*rG* [S.E.] = 0.28 [0.10]; GCP [S.E.] = -0.69 [0.21]) and sleep apnoea (*rG* [S.E.] = 0.32 [0.12]; GCP [S.E.] = -0.75 [0.17]). Military service (non-commissioned officer or other ranks) (*rG* [S.E.] = 0.57 [0.17]; GCP [S.E.] = -0.83 [0.13]) and traits related to trauma (e.g., 'been in serious accident believed to be life-threatening', *rG* [S.E.] = 0.33 [0.11]; GCP [S.E.] = -0.77 [0.16]) increased risk of chronic widespread pain.
