## Supplementary Figures for "A shared genetic signature for common chronic pain conditions and its impact on biopsychosocial traits"

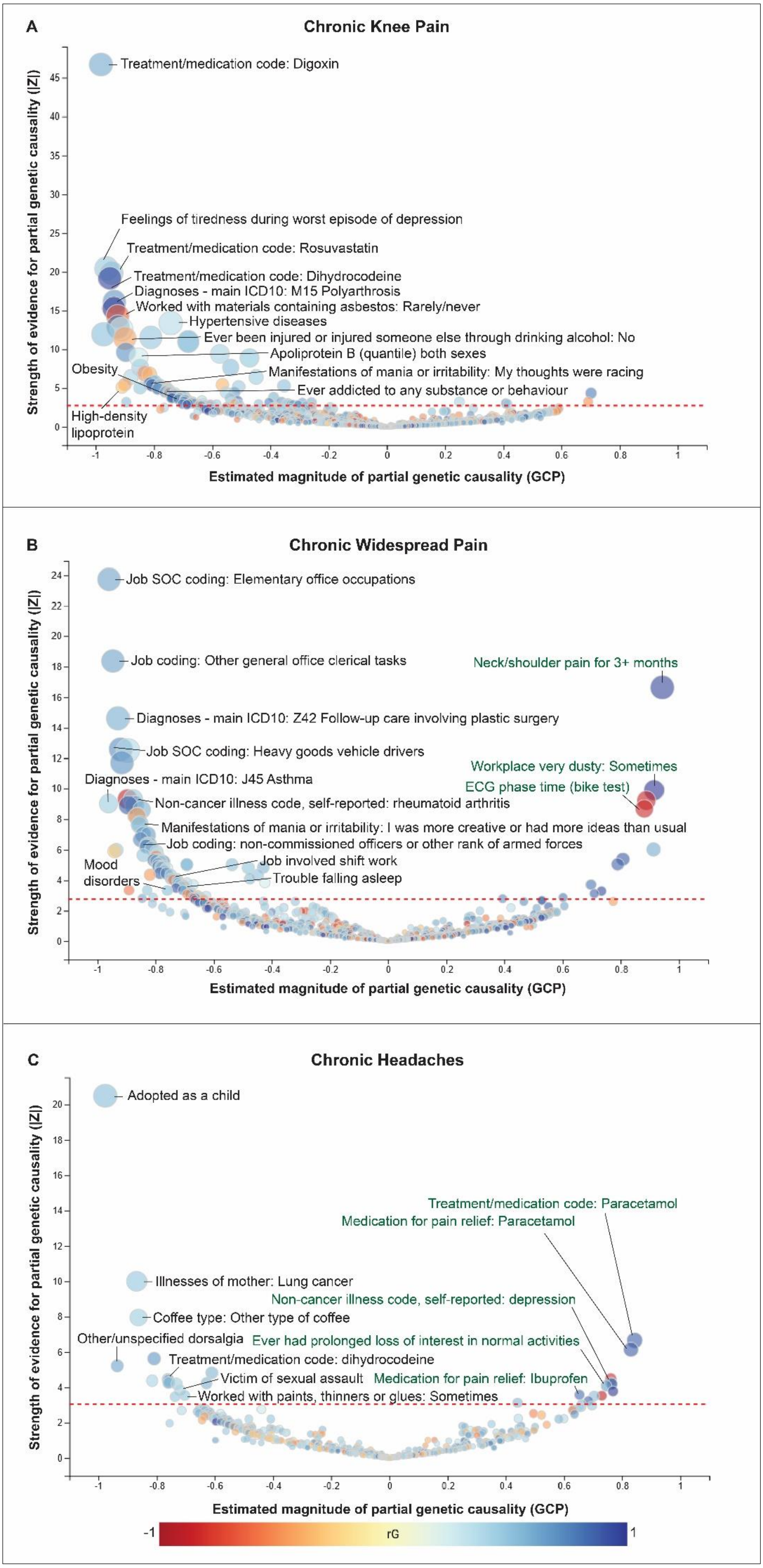

**Supplementary Figures:** Farrell et al. — 'A shared genetic signature for common chronic pain conditions and its impact on biopsychosocial traits'

**Supplementary Figure S1:** Causal architecture plots presenting the relationships of various traits with (**A**) chronic knee pain, (**B**) widespread pain and (**C**) headaches. Each dot is a data point representing a genetic correlation (*rG*). The colour of a dot indicates whether the genetic correlation is positive (red) or negative (blue). The x-axis denotes the genetic causal proportion (GCP) estimate and the y-axis denotes the GCP absolute Z-score (i.e., statistical significance). The red dashed line shows the threshold for statistical significance (5% false discovery rate). Traits that have a genetic causal effect on chronic pain are on the left side of the plot (with respect to 0 on the x-axis; GCP < -0.6, black text), while traits causally affected by chronic pain are on the right side (GCP > 0.6, green text). Selected traits are labelled for illustrative purposes — spanning clinical, biological, psychological and socioeconomic measures. Full details of all significant genetic causal relationships are provided in Supplementary Tables S5, S8 & S9.

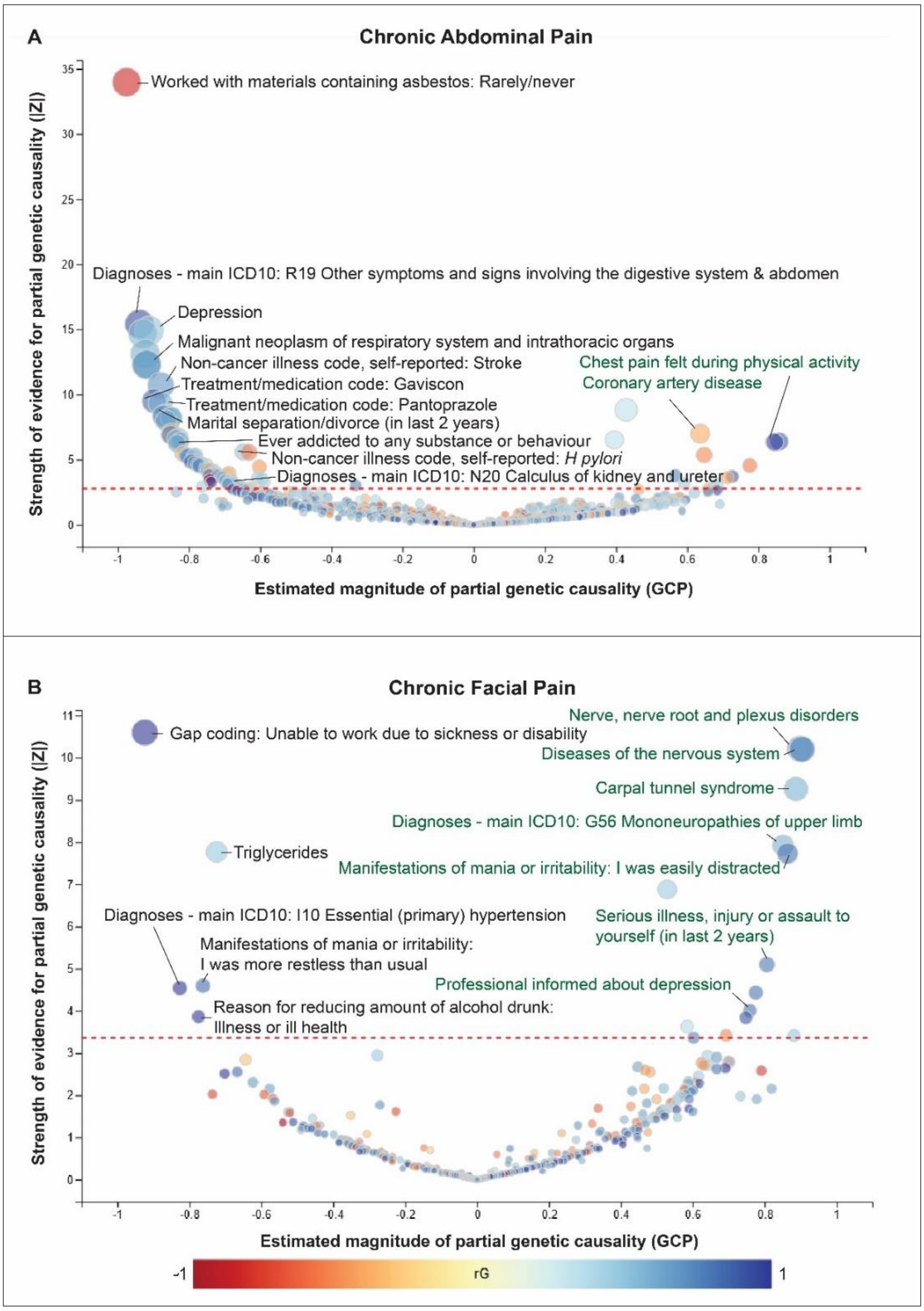


**Supplementary Figure S2:** Causal architecture plots presenting the relationships of various traits with (**A**) chronic abdominal pain and (**B**) facial pain. Each dot is a data point representing a genetic correlation (*rG*). The colour of a dot indicates whether the genetic correlation is positive (red) or negative (blue). The x-axis denotes the genetic causal proportion (GCP) estimate and the y-axis denotes the GCP absolute Z-score (i.e., statistical significance). The red dashed line shows the threshold for statistical significance (5% false discovery rate). Traits that have a genetic causal effect on chronic pain are on the left side of the plot (with respect to 0 on the x-axis; GCP < -0.6, black text), while traits causally affected by chronic pain are on the right side (GCP > 0.6, green text). Selected traits are labelled for illustrative purposes. Full details of all significant genetic causal relationships are provided in Supplementary Tables S6 & S7.
